# Prevalence, Incidence, and Mortality of Autoimmune Diseases Among Adolescents and Young Adults in Mexico: An Analysis Based on the Global Burden of Disease Study 2021

**DOI:** 10.1101/2025.04.04.25325208

**Authors:** Claudia Mendoza-Pinto, Pamela Munguía-Realpozo, Ivet Etchegaray-Morales, Fernanda Solis-Mendoza, José Luis Gálvez-Romero, Edith Ramírez-Lara, Marco Alejandro Trinidad González, Brenda Michel Silva Juárez, Máximo Alejandro García Flores, Socorro Méndez-Martínez

**Affiliations:** Departamento de Reumatología, Facultad de Medicina, Benemérita Universidad Autónoma de Puebla, Puebla, México; Unidad de Enfermedades Reumáticas y Autoinmunes Sistémicas, Unidad Médica de Alta Especialidad-Centro de Investigación Biomédica de Oriente, Instituto Mexicano del Seguro Social, Puebla, México; Departamento de Biología, Science School, Universidad de las Américas Puebla, San Andrés Cholula, Puebla, Mexico; Departamento de Investigación, Instituto de Seguridad y Servicios Sociales para los Trabajadores del Estado, Puebla; Coordinación de Educación e Investigación en Salud, Instituto Mexicano del Seguro Social, Puebla, Mexico

**Keywords:** Autoimmune diseases, Burden, Epidemiology, Adolescents and Young adults

## Abstract

Autoimmune diseases (ADs) pose a significant health burden among adolescents and young adults (AYAs) in Mexico. However, comprehensive national epidemiological data remain limited. This study evaluates the prevalence, incidence, and mortality of six major AD: rheumatoid arthritis (RA), inflammatory bowel disease (IBD), multiple sclerosis (MS), type 1 diabetes mellitus (T1DM), asthma, and psoriasis, utilizing data from the Global Burden of Disease Study 2021. Age-standardized rates were calculated through direct standardization using the GBD 2021 world standard population. Temporal trends from 1990 to 2021 were analyzed via Joinpoint regression, which determined each condition’s average annual percentage change (AAPC). Additionally, an ARIMA model was employed to forecast trends for the period from 2022 to 2035. In 2021, RA exhibited an age-standardized prevalence rate (ASPR) of 45.67, an incidence rate (ASIR) of 9.07, and a mortality rate (ASMR) of 0.02 per 100,000 inhabitants, with a marked female predominance and notable regional disparities. IBD showed low prevalence and incidence but a rising mortality trend, particularly in areas with enhanced diagnostic capacities. MS demonstrated increasing prevalence and incidence, especially among females. T1DM presented high prevalence and incidence, with declining trends over time accompanied by a gradual rise in mortality. Asthma remained highly prevalent despite overall declining trends, while psoriasis maintained a stable incidence with a considerable prevalence among AYAs. The findings highlight significant regional and sex-related disparities in the burden of ADs among Mexican AYAs. Forecasting analyses predict moderate increases in RA and MS, stability in IBD, asthma, and psoriasis, and a decline in T1DM prevalence and incidence, albeit with a slight increase in mortality. These results emphasize the urgent need for targeted public health interventions, improved diagnostic strategies, and equitable healthcare provision to mitigate the future impact of ADs in this vulnerable population.

## Introduction

Autoimmune diseases (ADs) are a diverse group of chronic disorders characterized by aberrant immune responses against self-antigens, leading to persistent inflammation and tissue damage ^1^. Globally, they affect approximately 7.6% to 9.4% of the population ^2^, with a significant burden among adolescents and young adults (AYAs) ^3^. These diseases include conditions such as rheumatoid arthritis (RA), systemic lupus erythematosus (SLE), inflammatory bowel disease (IBD), type 1 diabetes mellitus (T1DM), and multiple sclerosis (MS), all of which contribute to a substantial health burden ^4^. AYAs represent a unique demographic undergoing rapid physical, psychological, and social changes ^5^. The physiological processes of puberty and the transition to adulthood involve significant hormonal shifts and heightened stress levels, which can interact with immune dysregulation to exacerbate susceptibility to autoimmune conditions. For example, hormonal changes during puberty have been linked to the emergence and progression of ADs such as lupus and multiple sclerosis, conditions known for their higher prevalence in females during reproductive years. In males and females alike, ADs diagnosed in adolescence often lead to severe complications, including early cardiovascular disease, reduced bone density, and impaired psychosocial development ^6^.

The burden of ADs in AYAs is multifaceted. Beyond their immediate health impacts, these diseases impose long-term challenges on individuals’ quality of life ^7^. Chronic conditions such as RA and SLE not only result in physical disability but also hinder educational attainment, career development, and social integration during critical life stages ^8^. Furthermore, ADs often require complex, lifelong management strategies involving immunosuppressive treatments and regular monitoring, further complicating healthcare delivery and imposing significant economic costs.

Globally, significant progress has been achieved in elucidating the epidemiological dynamics of ADs, particularly in high-income countries ^9,10^. However, data on these conditions in low- and middle-income countries, including Mexico, remain sparse.

Mexico’s distinct demographic profile, characterized by an elevated prevalence of obesity, urbanization, and environmental risk factors, suggests potential variations in the epidemiology of ADs compared to other regions. Yet, comprehensive national-level data on the incidence, prevalence, and mortality of ADs among AYAs in Mexico are notably lacking. This gap poses a critical barrier to developing targeted prevention, diagnosis, and treatment strategies tailored to this vulnerable population. This study aims to address this gap by conducting a focused analysis of the prevalence, incidence, and mortality of ADs among Mexican AYAs. Through this research, we seek to provide a foundational understanding of the epidemiological profile of ADs in Mexico.

## Methods

### Data Source

The Global Burden of Disease Study (GBD) 2021 is recognized as the most extensive and comprehensive global epidemiological investigation conducted to date. It systematically evaluates the magnitude of health losses attributable to diseases, injuries, and risk factors across 204 countries and territories from 1990 to 2021 ^11^. The GBD database integrates data from diverse sources, including population-based surveys, cohort and registry studies, administrative health data, and other reports ^12^. The data are analyzed using DisMod-MR 2.1, a state-transition disease modeling tool for epidemiologic analyses, alongside MR-BRT, a Bayesian meta-regression software, to ensure coherent estimates of disease burden across locations and time ^13^.

For each condition, the GBD uses 1000 draw-level estimates to capture the uncertainty in model predictions, defining 95% uncertainty intervals (UIs) as the range between the 2.5th and 97.5th percentiles of these draws ^14^. This study used the GBD results tool (http://ghdx.healthdata.org/gbd-results-tool) to extract estimates and UIs (per 100,000 population). Variables analyzed included incidence, prevalence, and mortality, categorized by age (15–24 years), sex, calendar year (1990–2021), and geographic location (national and state levels). Specific ADs assessed in this study included RA, IBD, MS, T1DM, asthma, and psoriasis.

### Case Definition of Autoimmune Diseases

The diagnosis and classification of ADs in this study adhered to clinical guidelines established by the World Health Organization (WHO) and the standards set by the International Statistical Classification of Diseases and Related Health Problems, 10th Revision (ICD-10) (https://icd.who.int/browse10/2019/en). Each condition was categorized using its corresponding ICD-10 codes. RA was coded as M05–M05.9 and M08–M09.8, while IBD was identified under K50 (Crohn’s disease), K51 (ulcerative colitis), K52 (indeterminate colitis), K52.8 (other specified noninfective gastroenteritis and colitis), and K52.9 (noninfective gastroenteritis and colitis, unspecified). MS was classified as G35–G35.0, and T1DM was captured under E10–E10.11 and E10.3–E10.9. Asthma was represented by codes J45–J46.0, and psoriasis fell within the range L40–L41.9. These standardized classifications ensured consistency in identifying and analyzing the targeted diseases.

### Statistical analysis

Age-standardized rates (ASRs) were derived utilizing the direct standardization method and applying weights derived from the GBD 2021 world standard population. These rates were expressed per 100,000 individuals alongside their respective 95% uncertainty intervals (UIs) ^12,15^. To evaluate trends in age-specific incidence (ASIR), prevalence (ASPR), and mortality (ASMR) rates of ADs on global, continental, and national scales, Joinpoint regression analysis was utilized^16^.

Joinpoint regression software (version 5.1.0, accessible at https://surveillance.cancer.gov/joinpoint/) enabled trend analysis from 1990 to 2021. This analysis computed the average annual percentage change (AAPC) and its 95% confidence intervals (CIs). Trends were classified as increasing (AAPC > 0), decreasing (AAPC < 0), or stable (AAPC 95% CI encompassing 0). It is crucial to distinguish that CIs primarily reflect sampling error uncertainty, whereas UIs incorporate uncertainty from various sources, including sampling, model estimations, and specifications ^17^.

An ARIMA model is a type of differential, integral, moving average, and autoregressive model, also known as an “integral moving average autoregressive model”, and represents a model commonly used to use time series of data for forecasting analysis. In the ARIMA model (*p, d, q*), AR is “autoregressive”, *p* is the number of autoregressive terms, MA is “moving average”, *q* is the number of terms in the moving average, and d is the number of differences (orders) that provide a smooth series ^18^. The optimal model was automatically filtered by the Akaike information criterion (AIC) and Bayesian information criterion (BIC). In this study, the ARIMA model was used to analyze the burden of disease based on the trend for disease burden and to predict the burden of disease associated with ADs in AYAs n Mexico between 2022 and 2035.

## Results

### Rheumatoid Arthritis

In 2021, RA affected AYAs in Mexico with global ASPR, ASIR , and ASMR of 45.67 (95% UI: 30.34-66.35), 9.07 (95% UI: 6.29-12.61), and 0.02 (95% UI: 0.02-0.03) per 100,000 inhabitants, respectively (Table 1). Women were disproportionately affected, showing higher values across all indicators compared to men. Significant geographic disparities were noted; Oaxaca exhibited the highest ASPR and ASIR (53.23; 95% UI: 34.69-78.76 and 11.14; 95% UI: 7.80-15.57, respectively), while Zacatecas recorded the highest mortality rate (0.05; 95% UI: 0.04-0.07). Conversely, Baja California, Baja California Sur, and Nuevo Leon reported the lowest figures for all three metrics. Trends from 1990 to 2021 revealed a marked increase in ASPR and ASIR, paired with a significant decline in mortality (Table 3).

**Table 1.**
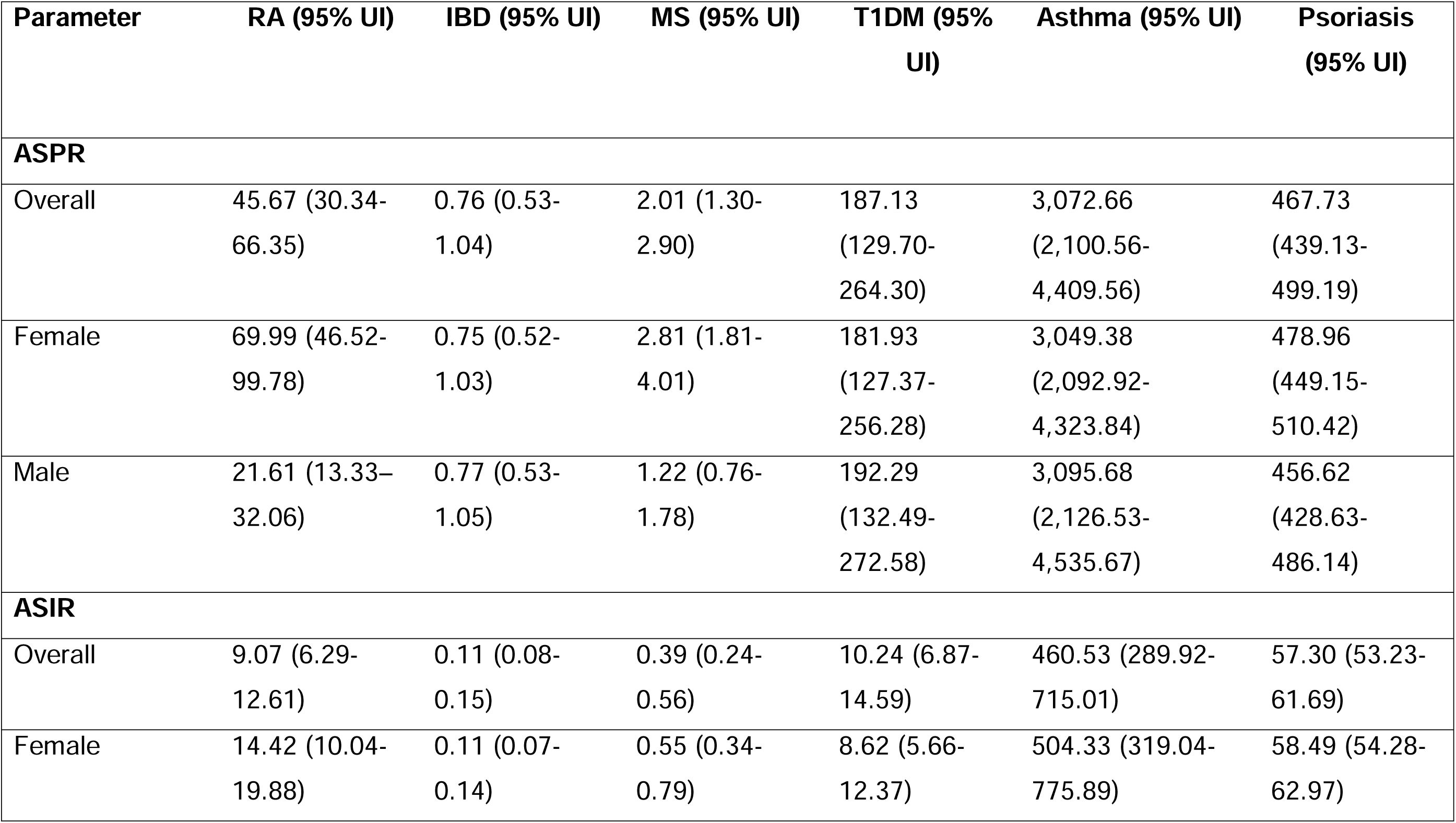
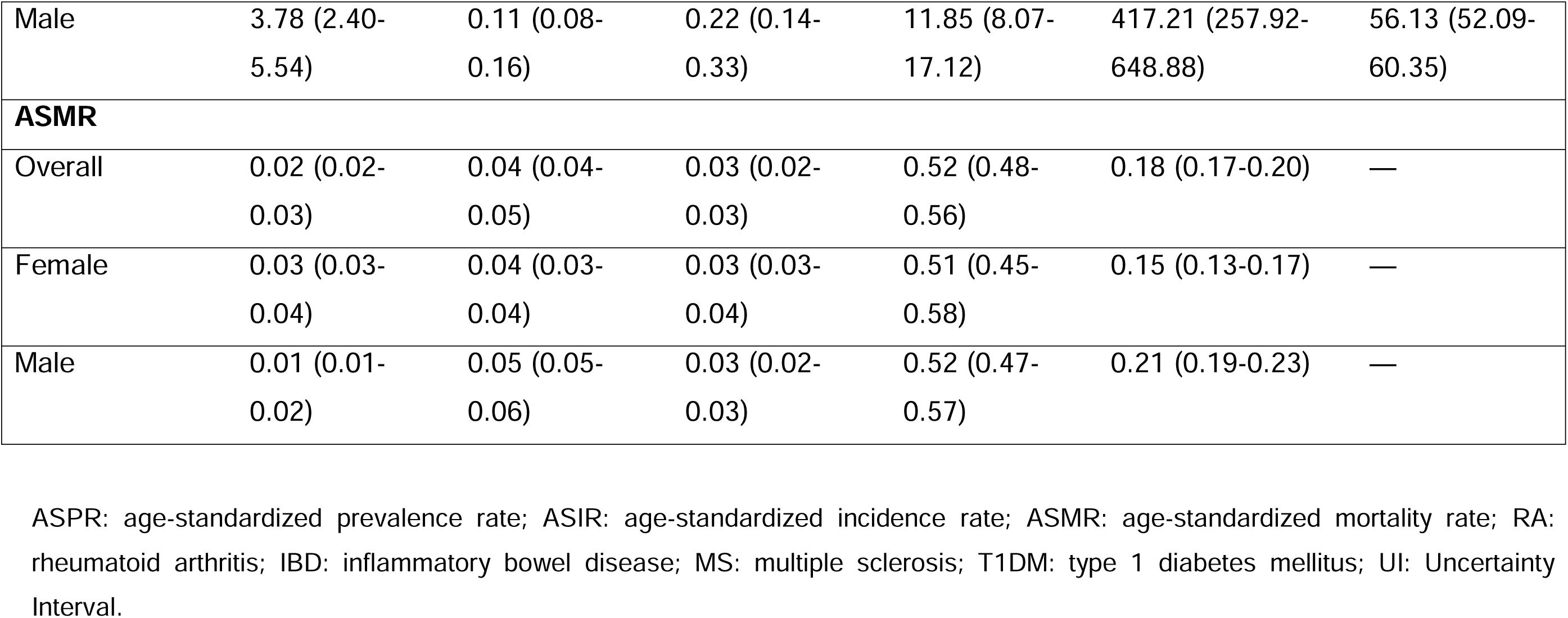
ASPR, ASIR, and ASMR of autoimmune diseases in adolescents and young in Mexico in 2021.

| Parameter | RA (95% UI) | IBD (95% UI) | MS (95% UI) | T1DM (95% UI) | Asthma (95% UI) | Psoriasis (95% UI) |
| --- | --- | --- | --- | --- | --- | --- |
| **ASPR** |  |  |  |  |  |  |
| Overall | 45.67 (30.34-66.35) | 0.76 (0.53-1.04) | 2.01 (1.30-2.90) | 187.13 (129.70-264.30) | 3,072.66 (2,100.56-4,409.56) | 467.73 (439.13-499.19) |
| Female | 69.99 (46.52-99.78) | 0.75 (0.52-1.03) | 2.81 (1.81-4.01) | 181.93 (127.37-256.28) | 3,049.38 (2,092.92-4,323.84) | 478.96 (449.15-510.42) |
| Male | 21.61 (13.33-32.06) | 0.77 (0.53-1.05) | 1.22 (0.76-1.78) | 192.29 (132.49-272.58) | 3,095.68 (2,126.53-4,535.67) | 456.62 (428.63-486.14) |
| **ASIR** |  |  |  |  |  |  |
| Overall | 9.07 (6.29-12.61) | 0.11 (0.08-0.15) | 0.39 (0.24-0.56) | 10.24 (6.87-14.59) | 460.53 (289.92-715.01) | 57.30 (53.23-61.69) |
| Female | 14.42 (10.04-19.88) | 0.11 (0.07-0.14) | 0.55 (0.34-0.79) | 8.62 (5.66-12.37) | 504.33 (319.04-775.89) | 58.49 (54.28-62.97) |
| Male | 3.78 (2.40-5.54) | 0.11 (0.08-0.16) | 0.22 (0.14-0.33) | 11.85 (8.07-17.12) | 417.21 (257.92-648.88) | 56.13 (52.09-60.35) |
| **ASMR** |  |  |  |  |  |  |
| Overall | 0.02 (0.02-0.03) | 0.04 (0.04-0.05) | 0.03 (0.02-0.03) | 0.52 (0.48-0.56) | 0.18 (0.17-0.20) | — |
| Female | 0.03 (0.03-0.04) | 0.04 (0.03-0.04) | 0.03 (0.03-0.04) | 0.51 (0.45-0.58) | 0.15 (0.13-0.17) | — |
| Male | 0.01 (0.01-0.02) | 0.05 (0.05-0.06) | 0.03 (0.02-0.03) | 0.52 (0.47-0.57) | 0.21 (0.19-0.23) | — |

ASPR: age-standardized prevalence rate; ASIR: age-standardized incidence rate; ASMR: age-standardized mortality rate; RA: rheumatoid arthritis; IBD: inflammatory bowel disease; MS: multiple sclerosis; T1DM: type 1 diabetes mellitus; UI: Uncertainty Interval.

**Table 2.**
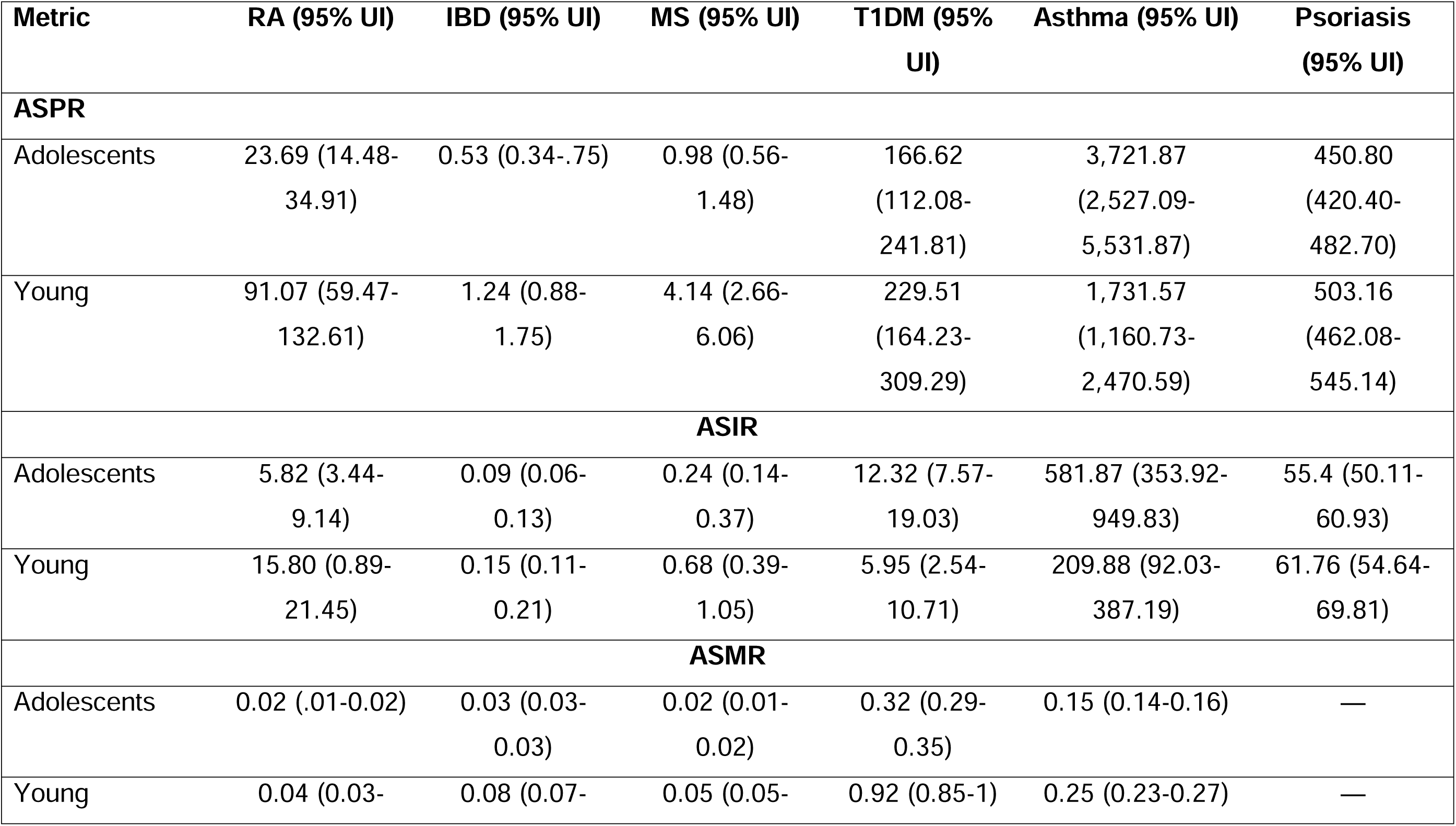
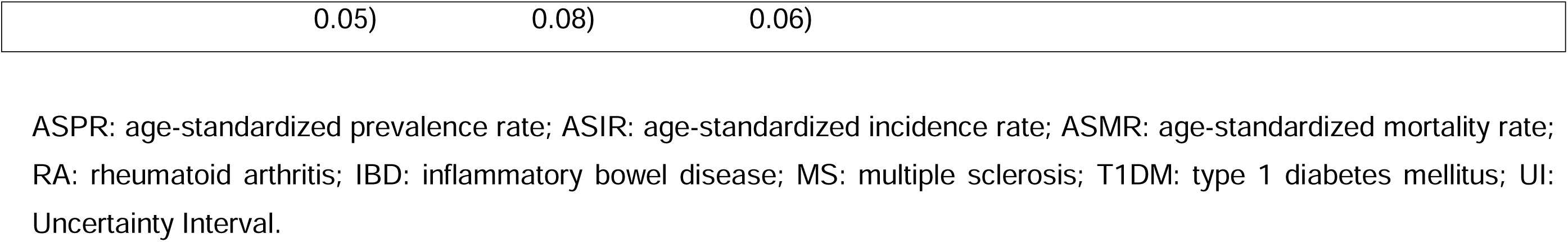
ASPR, ASIR, and ASMR of autoimmune diseases in adolescents (10-19 years) and young adults (20-24 years) in Mexico in 2021.

**Table 3.** Changing trends of ASPR, ASIR, and ASMR of autoimmune diseases in adolescents and young adults in Mexico from 1990 to 2021.

|  | Average Annual Percent Change % (95% CI) |  |  |  |  |  |
| --- | --- | --- | --- | --- | --- | --- |
|  | RA | IBD | MS | T1DM | Asthma | Psoriasis |
| **ASPR** | 0.90 (0.86-0.93)* | -0.20 (-0.05-0.01) | 1.30 (1.20-1.40)* | -0.60 (-0.50- -<br>0.070)* | -1.10 (-1.30- -<br>0.80)* | 0.30 (0.20-0.40)* |
| **ASIR** | 0.90 (0.84-0.92)* | -0.05 (-0.90- -<br>0.10)** | 1.50 (1.30-1.60)* | -0.11 (-0.10- -<br>0.08)* | -0.70 (-0.90- -<br>0.60)* | 0.30 (0.20-0.30)* |
| **ASMR** | -2.20 (-1.70- -<br>2.60)* | 0.80 (0.50-1.10)* | 4.10 (2.50-5.70)* | 1.30 (0.80-1.90)* | -1.80 (-3.10- -<br>0.40)* | — |

ASPR: age-standardized prevalence rate; ASIR: age-standardized incidence rate; ASMR: age-standardized mortality rate; RA: rheumatoid arthritis; IBD: inflammatory bowel disease; MS: multiple sclerosis; T1DM: type 1 diabetes mellitus; CI: Confidence Interval.

### Inflammatory Bowel Disease

The burden of IBD in AYAs in Mexico during 2021 was characterized by ASPR, ASIR and ASMR of 0.76 (95% UI: 0.53-1.04), 0.11 (95% UI: 0.08-0.15), and 0.04 (95% UI: 0.04-0.05) per 100,000 inhabitants, respectively. Unlike RA, no significant sex differences were observed. Mexico City emerged as the region with the highest prevalence (0.85; 95% UI: 0.59-1.17) and incidence (0.12; 95% UI: 0.08-0.16), while Zacatecas led in mortality (0.08; 95% UI: 0.07-0.10). At the opposite end, Chiapas, Nayarit, and Campeche registered the lowest metrics. Over the 1990-2021 period, prevalence remained stable, while incidence exhibited a downward trend and mortality increased significantly (Table 3).

### Multiple Sclerosis

MS presented a global ASPR of 2.01 (95% UI: 1.30-2.90), ASIR 0.39 (95% UI: 0.24-0.56), and ASMR of 0.03 (95% UI: 0.02-0.03) per 100,000 inhabitants in Mexico for 2021. Women consistently demonstrated higher rates than men. Baja California led in both prevalence (2.62; 95% UI: 1.68-3.79) and incidence (0.51; 95% UI: 0.32-0.75), while Zacatecas reported the highest mortality rate (0.07; 95% UI: 0.06-0.08). Trends over time highlighted increasing rates across all metrics, underscoring a growing burden of MS (Table 3).

### Type 1 Diabetes Mellitus

In 2021, T1DM in Mexican AYAs showed ASPR, ASIR and ASMR of 187.13 (95% UI: 129.70-264.30), 10.24 (95% UI: 6.87-14.59), and 0.52 (95% UI: 0.48-0.56) per 100,000 inhabitants, respectively. Males were slightly more affected than females. Mexico City, Chiapas, and Guanajuato exhibited the highest prevalence (197.64; 95% UI: 137.55-279.67), incidence (10.33; 95% UI: 7.12-15.04), and mortality (0.82; 95% UI: 0.73-0.92), respectively. The lowest values were observed in Hidalgo, Nuevo Leon, and Yucatan. Longitudinal analysis from 1990 to 2021 revealed a decline in ASPR and ASIR, in contrast to an upward trend in mortality (Table 3).

### Asthma

Asthma ASPR, ASIR and ASMR in AYAs in Mexico for 2021 were 3,072.66 (95% UI: 2,100.56-4,409.56), 460.53 (95% UI: 289.92-715.01), and 0.18 (95% UI: 0.17-0.20) per 100,000 inhabitants, respectively. While ASPRs were comparable between sexes, females exhibited higher incidence and mortality. Veracruz registered the highest prevalence (3,561.27; 95% UI: 2,459.70-4,561.27) and incidence (524.84; 95% UI: 336.90-807.38), while Tabasco had the highest mortality (0.33; 95% UI: 0.27-0.40). Mexico City and Nuevo Leon recorded the lowest values across all metrics. Asthma trends between 1990 and 2021 demonstrated consistent declines in ASPR, ASIR, and ASMR (Table 3).

### Psoriasis

Psoriasis affected Mexican AYAs in 2021 with ASPR and ASIR of 467.73 (95% UI: 439.13-499.19) and 57.30 (95% UI: 53.23-61.69) per 100,000 inhabitants, respectively. Females were more impacted than males. Mexico City reported the highest rates (508.14; 95% UI: 473.74-544.61 and 60.25; 95% UI: 55.49-65.79 for prevalence and incidence, respectively), while Chiapas registered the lowest values. No mortality data were available for psoriasis in the GBD study. Analysis from 1990 to 2021 indicated a gradual decline in ASPR and ASIR (Table 3).

### Forecasted Trends by Disease (2022–2035)

#### Rheumatoid Arthritis (RA)

Projections indicate a gradual increase in the ASPR (Fig. 1a) and ASIR of RA, rising from 46.08 (95% CI: 45.93–46.22) to 51.44 (95% CI: 42.97–59.92) and from 9.14 (95% CI: 9.10–9.17) to 10.64 (95% CI: 8.12–13.17) per 100,000 inhabitants, respectively. The ASMR is expected to remain stable at 0.02, with marginal variation (Table S1).

**Figure 1.**
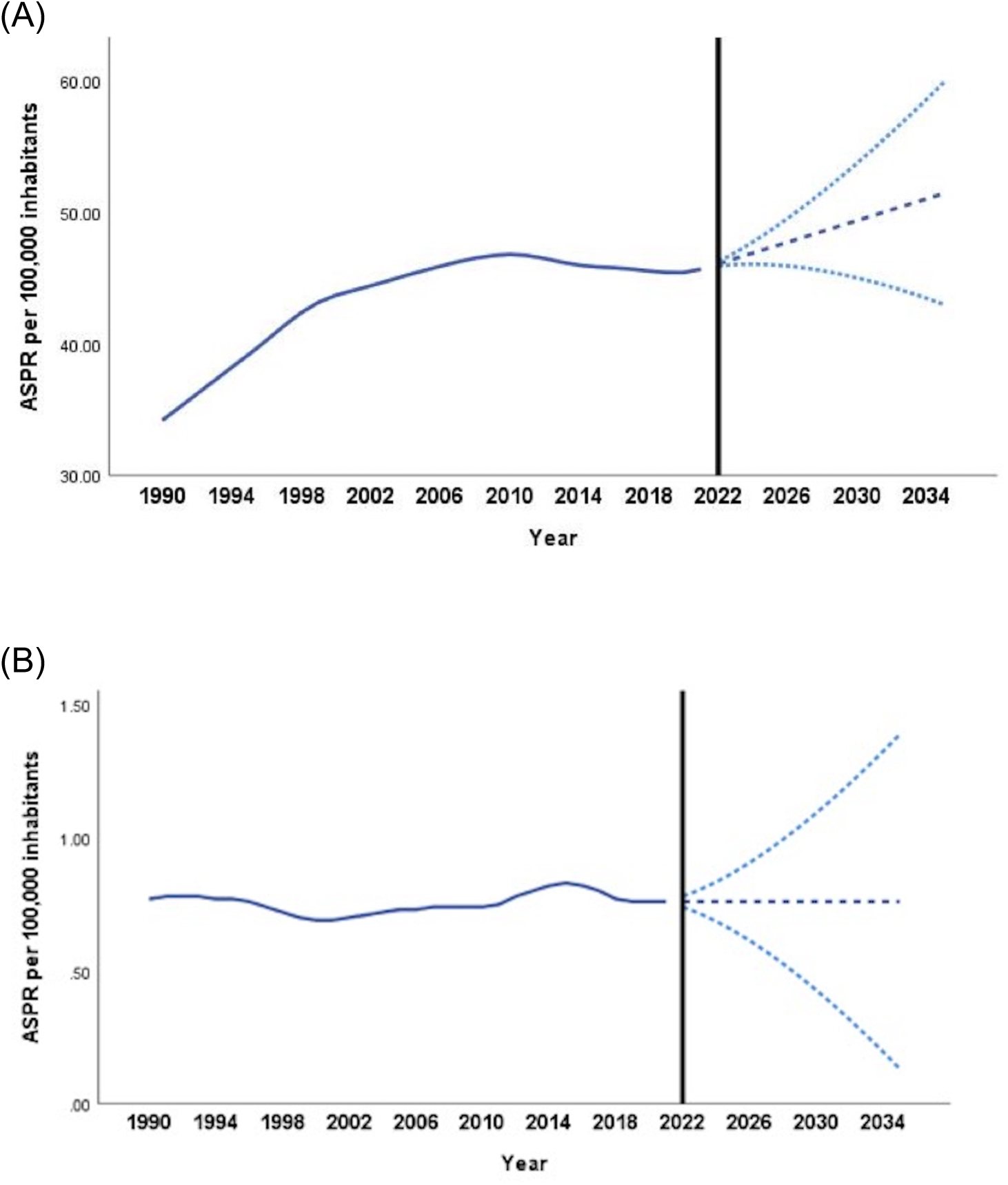
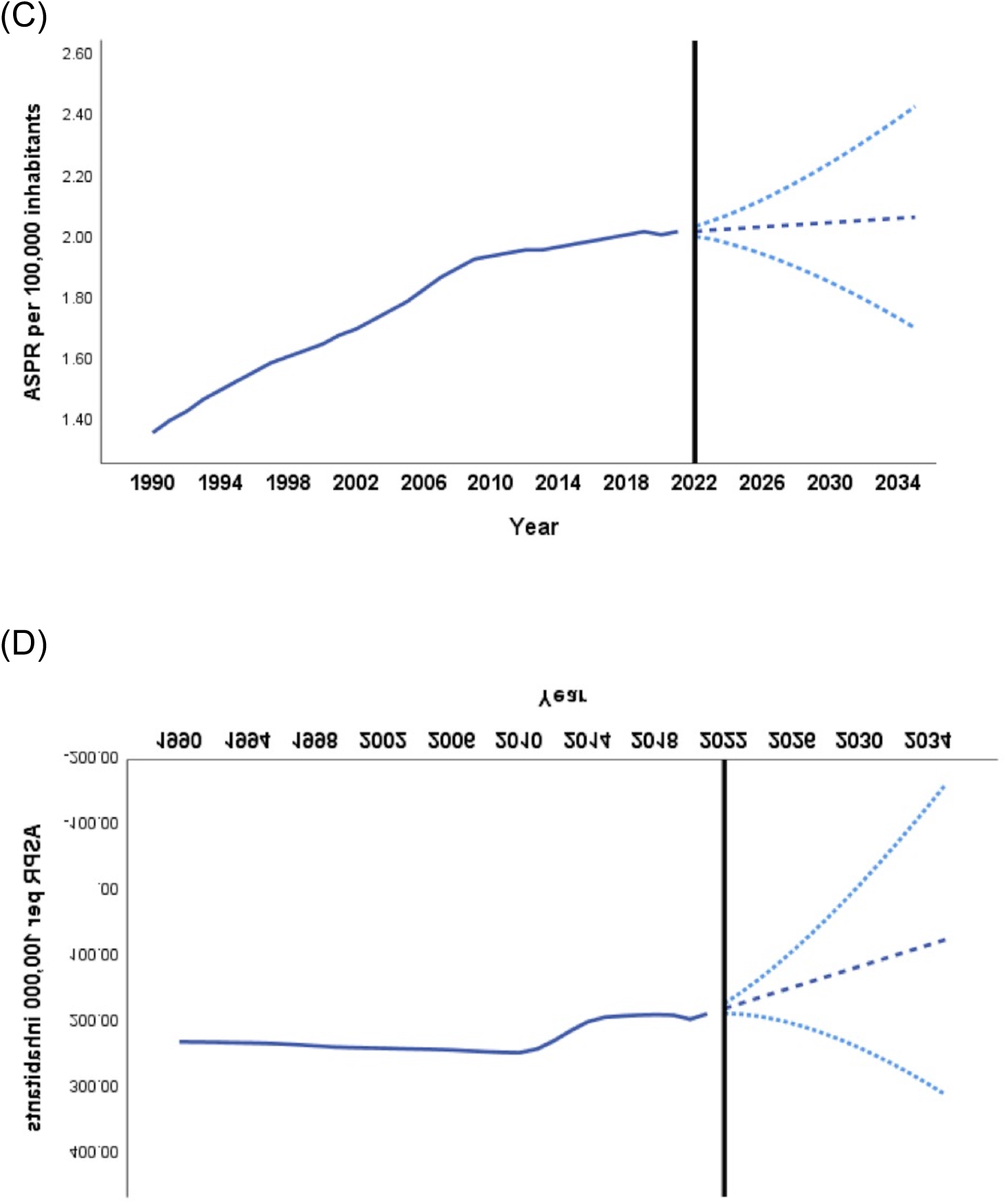
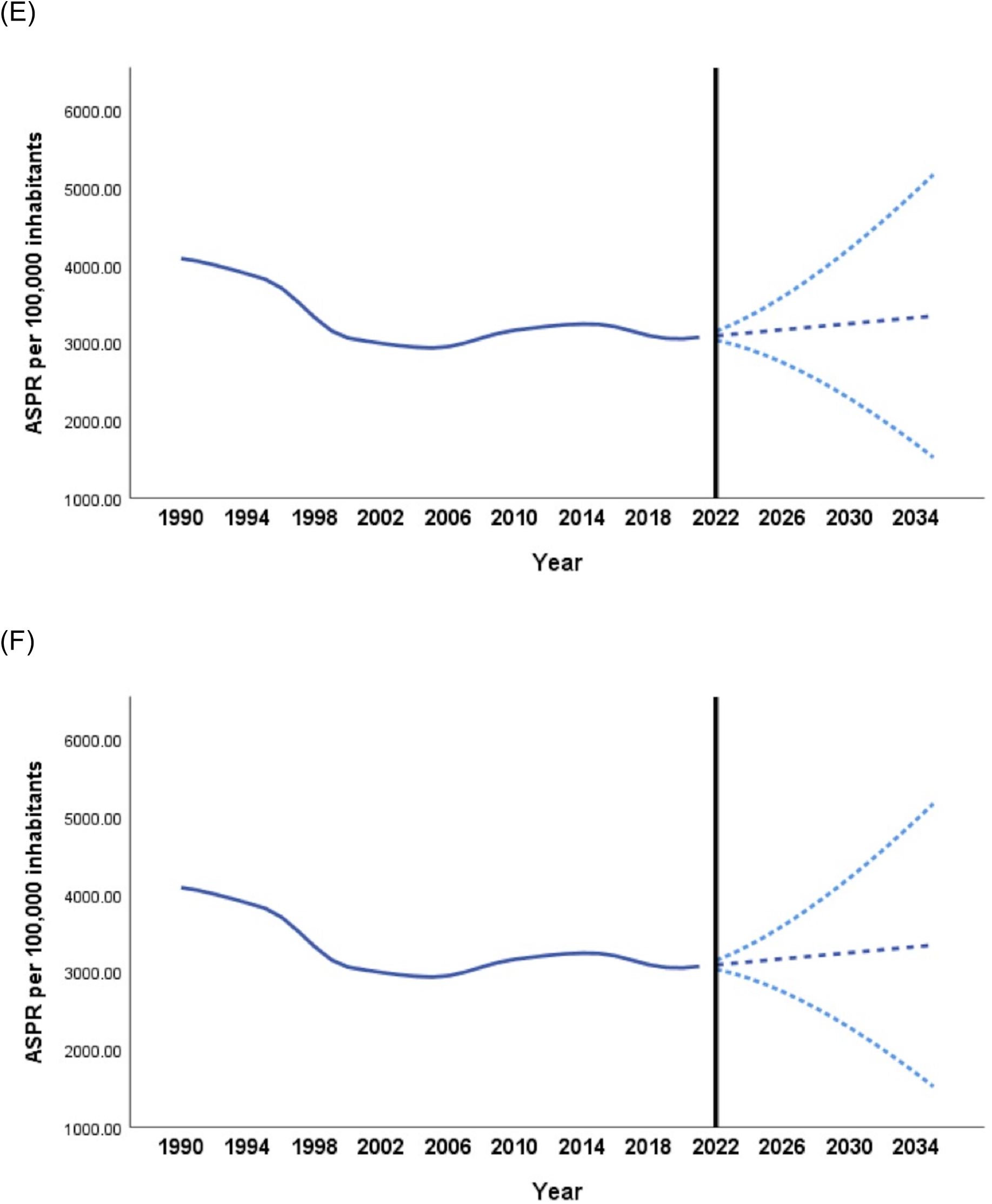
**A-F.** Projected trends in the age-standardized prevalence rate (ASPR) for six autoimmune and chronic inflammatory diseases (1990–2035) in adolescents and young adults in Mexico. The solid lines represent observed trends (1990–2021), and the dashed lines indicate projected rates (2022–2035) with 95% confidence intervals: (A) rheumatoid arthritis, (B) inflammatory bowel disease, (C) multiple sclerosis, (D) type 1 diabetes mellitus, (E) asthma, and (F) psoriasis.

### Inflammatory Bowel Disease (IBD)

The ASPR of IBD is projected to remain constant at 0.76 (95% CI: 0.74–0.78 in 2022; 0.56–0.96 in 2035) per 100,000 inhabitants (Fig. 1b). Both ASIR and ASMR exhibit no significant changes, maintaining rates of approximately 0.11 and 0.04, respectively (Table S2).

### Multiple Sclerosis (MS)

The forecast reveals minor increases in ASPR (Fig. 1c) and ASIR, with prevalence rising from 2.02 (95% CI: 2.00–2.03) in 2022 to 2.07 (95% CI: 1.76–2.39) per 100,000 in 2035. Mortality is predicted to remain low, increasing slightly from 0.03 to 0.04 per 100,000 inhabitants (Table S3).

### Type 1 Diabetes Mellitus (T1DM)

Projected ASPR demonstrates a significant decline, decreasing from 179.04 (95% CI: 171.62–186.46) in 2022 to 73.87 (95% CI: -162.62–310.36) per 100,000 in 2035 (Fig. 1d). Similarly, ASIR declines from 10.03 to 7.28, while ASMR gradually increases from 0.52 to 0.58 during the same period (Table S4).

### Asthma

While asthma’s ASPR is projected to increase slightly from 3,092.27 (95% CI: 3,034.98–3,149.56) in 2022 to 3,347.19 (95% CI: 1,522.09–5,172.29) per 100,000 inhabitants by 2035 (Fig. 1e), both ASIR and ASMR exhibit minimal changes, maintaining rates near 460 and 0.12 per 100,000, respectively (Table S5).

### Psoriasis

The projections show negligible changes in both ASPR and ASIR for psoriasis, with prevalence expected to remain at 467.71 (95% CI: 467.12–468.30) in 2022 and 467.45 (95% CI: 448.75–486.15) in 2035 (Fig. 1f). Similarly, incidence rates remain stable (Table S6).

## Discussion

This study presents the first comprehensive assessment of the burden posed by six major autoimmune diseases—RA, IBD, MS, T1DM, asthma, and psoriasis— among AYAs in Mexico. Drawing on data from the GBD 2021 project, we examined ASPR, ASIR, and ASMR at both the global and regional levels, encompassing trends from 1990 to 2021. Furthermore, a forecasting analysis estimated the trajectory of these diseases through 2035, offering critical insight into the likely future challenges in healthcare resource allocation and disease management.

Our findings underscore a notable and multifaceted disease burden of RA in the AYA population. In 2021, the ASPR, ASIR, and ASMR of RA were significantly higher in females, corroborating well-established global epidemiological patterns ^19^. Of particular interest, Oaxaca displayed the highest ASPR and ASIR, while Zacatecas recorded the highest mortality. These disparities may reflect regional differences in healthcare access, socioeconomic resources, or the timeliness of diagnosis and treatment strategies ^20^. Over the 1990–2021 interval, a marked rise in both prevalence and incidence resonates with growing disease awareness, improved diagnostic proficiency, and potentially evolving environmental factors.

Encouragingly, RA-related mortality declined, likely indicative of more effective management protocols, including earlier therapeutic intervention and the advent of biologic therapies ^21,22^. These observations echo prior reports of a significant reduction in RA mortality rates in Mexico after 2004 ^23^. Altogether, our results highlight the continued need for region-specific prevention and treatment strategies, particularly in underserved areas, to counteract the rising RA burden and to ensure equitable healthcare provision for younger populations.

In contrast, IBD exhibited a comparatively low prevalence, incidence, and mortality among AYAs, with no clear sex predisposition. Interestingly, these results diverge from other Latin American data showing a rising IBD burden, especially in pediatric populations ^24^. In our study, Mexico City displayed the highest prevalence and incidence, possibly due to greater availability of specialized care and better diagnostic infrastructure ^25^. However, the overall static prevalence trend and declining incidence from 1990 to 2021 may point to underdiagnosis or underreporting in less-resourced areas. Despite stable or decreasing incidence rates, the rising IBD mortality is concerning and highlights ongoing challenges in managing severe disease and complications in youth ^26,27^. These findings reinforce the need to bolster access to specialty care, address disparities in diagnosis and treatment, and ensure timely intervention across all regions of Mexico.

Our analysis also sheds light on the epidemiological landscape of MS among AYAs. As reported in the “Registro Mexicano de Esclerosis Multiple” (REMEMBer) study ^28^, MS displayed a pronounced female predominance, a pattern mirrored globally ^29^. However, unlike high-income regions where MS incidence and prevalence have been increasing, our data suggest relatively low rates in Mexico, raising the possibility of underdiagnosis or limited access to specialized neurologic care. These observations highlight the need for systematic screening and improved resource allocation to neurology services, particularly in regions where diagnostic and treatment facilities remain insufficient.

T1DM emerged as another condition with relatively low documented prevalence, incidence, and mortality rates among Mexican AYAs. These findings may indicate a genuinely lower burden of T1DM or, alternatively, reinforce potential underdiagnosis—particularly in rural or underserved areas. National registry data spanning 2000–2018 ^30^ further nuance this picture, illustrating temporal fluctuations in T1DM incidence, including increases between 2000 and 2006, decreases thereafter, and potential spikes associated with environmental events such as influenza outbreaks. Elevated incidence in older pediatric cohorts (10–19 years) and higher rates among females highlight the complex interplay of genetic predisposition, environmental triggers, and possibly differential healthcare-seeking behaviors. These dynamics stress the necessity for robust diabetes registries and systematic public health interventions to detect T1DM early and manage it effectively in AYAs.

Asthma remains an important respiratory condition in this demographic, with a prevalence of 3,072.66 per 100,000, an incidence of 460.53 per 100,000, and a mortality rate of 0.18 per 100,000 in 2021. Although our findings demonstrate a decline in prevalence, incidence, and mortality from 1990 to 2021, disparities persist, most notably the higher mortality rate observed in females. Geographically, Veracruz had the highest prevalence and incidence, whereas Tabasco registered the highest mortality. By contrast, urbanized regions such as Mexico City and Nuevo León exhibited the lowest metrics across these domains. These patterns parallel observations in pediatric cohorts (0–14 years), suggesting that improvements in early detection, standardized management protocols, and strengthen public health policies may have contributed to better outcomes overall but have not entirely mitigated rural-urban and sex-based inequalities ^31,32^. Future interventions must continue to prioritize equitable healthcare access, environmental risk reduction, and awareness campaigns to further reduce the asthma burden.

Psoriasis also warrants particular attention due to its substantial burden in adolescence and early adulthood. Beyond the hallmark cutaneous manifestations, young individuals with psoriasis face increased risks of developing comorbidities such as metabolic syndrome, cardiovascular disease ^33^, and psychosocial conditions including depression and anxiety ^34^. Our findings stress the necessity to elucidate the potential sex-related discrepancies in psoriasis incidence and prevalence and to determine whether underlying biological mechanisms, healthcare utilization patterns, or differences in health-seeking behavior contribute to these trends. Prospective research should explore the longitudinal progression of psoriasis severity and comorbidities, as well as integrate cost-effective, region-specific interventions that effectively reduce disease burden and enhance quality of life in young adults.

Forecasting analysis offers valuable insights into the shifting epidemiological profile of these six autoimmune and chronic inflammatory conditions. Our projections suggest that RA and MS may experience moderate increases in prevalence and incidence, while conditions like asthma and psoriasis remain largely stable.

Notably, T1DM is predicted to decline in prevalence and incidence, contrasted with a slight increase in mortality rates. IBD, meanwhile, is expected to remain relatively rare, displaying minimal alterations over time. These diverse trajectories bring out the complex challenges of managing autoimmune diseases in AYAs. The persistent or increasing prevalence of certain conditions demands sustained focus on timely detection, improved accessibility to specialized care, and equitable treatment regimens, whereas the declining incidence of T1DM should not divert attention from the modest but concerning rise in mortality.

Despite offering a comprehensive overview, this study has certain limitations. First, reliance on GBD data—strongly influenced by modeling techniques—may introduce biases stemming from variations in data quality across countries.

Inconsistencies in disease reporting and underdiagnosis in low-resource settings can skew prevalence and incidence estimates, while well-resourced regions might inflate disease burden estimates due to advanced diagnostic capabilities. Second, the absence of subgroup analyses precludes a finer-grained understanding of disease patterns in vulnerable populations; subdividing by socioeconomic status, narrower age brackets, or high-risk cohorts might elucidate distinct risk factors, informing more effective public health measures. Lastly, heterogeneity in healthcare infrastructure and surveillance methods across regions may limit comparability and accentuate the risk of data misinterpretation. Recognizing these caveats, further research should aim to refine epidemiological estimates through rigorous primary data collection, systematic subgroup analyses, and harmonized reporting standards.

In conclusion, this study provides a nationwide perspective on the epidemiological burden of six key autoimmune and chronic inflammatory diseases— RA, IBD, MS, T1DM, asthma, and psoriasis—among adolescents and early adults in Mexico. The results reveal significant regional disparities, sex-based differences in disease burden, and varying trends over time. RA and MS exhibit rising prevalence and incidence, whereas T1DM shows declining incidence and prevalence but increasing mortality. Asthma’s gradual decline in rates contrasts with IBD’s relatively stable burden yet concerning rise in mortality. Psoriasis contributes a substantial non-fatal burden but lacks direct mortality data in the GBD dataset. These findings indicate the need for targeted interventions, improved surveillance, and equitable healthcare access to mitigate the future impact of these conditions on Mexico’s younger population.

## Data Availability

All data produced in the present study are available upon reasonable request to the authors

## Conflict of interest

The authors declare no potential conflicts of interest.

## Author contributions

Conception and design of the study: CMP, PMR, IEM; Acquisition of data: All; Analysis and interpretation of data: FSP, JLGR, ERL, MATG, BMSJ; Drafting of the manuscript or revising it for important content: CMP, PMR, MAGF, SMM; Final approval of the version submitted for publication: All.

## Notes

### Competing Interest Statement

The authors have declared no competing interest.

### Funding Statement

This study did not receive any funding

## References

1. Miller FW. The increasing prevalence of autoimmunity and autoimmune diseases: an urgent call to action for improved understanding, diagnosis, treatment, and prevention. Curr Opin Immunol. 2023;80:102266. doi:10.1016/j.coi.2022.102266

2. Gutierrez-Arcelus M, Rich SS, Raychaudhuri S. Autoimmune diseases - connecting risk alleles with molecular traits of the immune system. Nat Rev Genet. 2016;17:160–174. doi:10.1038/nrg.2015.33

3. Zhao M, Zhai H, Li H, et al. Age-standardized incidence, prevalence, and mortality rates of autoimmune diseases in adolescents and young adults (15-39 years): an analysis based on the global burden of disease study 2021. BMC Public Health. 2024;24:1800. doi:10.1186/s12889-024-19290-3

4. Cao F, He Y-S, Wang Y, et al. Global burden and cross-country inequalities in autoimmune diseases from 1990 to 2019. Autoimmun Rev. 2023;22:103326. doi:10.1016/j.autrev.2023.103326

5. Tunnicliffe DJ, Singh-Grewal D, Chaitow J, et al. Lupus Means Sacrifices: Perspectives of Adolescents and Young Adults With Systemic Lupus Erythematosus. Arthritis Care Res (Hoboken). 2016;68:828–837. doi:10.1002/acr.22749

6. Chan P-C, Yu C-H, Yeh K-W, Horng J-T, Huang J-L. Comorbidities of pediatric systemic lupus erythematosus: A 6-year nationwide population-based study. J Microbiol Immunol Infect. 2016;49:257–263. doi:10.1016/j.jmii.2014.05.001

7. Zigler CK, Li Z, Hernandez A, et al. Evaluating anchor variables and variation in meaningful score differences for PROMIS(®) Pediatric measures in children and adolescents living with a rheumatic disease. Qual life Res. 2024;33:3449–3457. doi:10.1007/s11136-024-03800-2

8. Malviya A, Rushton SP, Foster HE, et al. The relationships between adult juvenile idiopathic arthritis and employment. Arthritis Rheum. 2012;64:3016–3024. doi:10.1002/art.34499

9. Conrad N, Misra S, Verbakel JY, et al. Incidence, prevalence, and co-occurrence of autoimmune disorders over time and by age, sex, and socioeconomic status: a population-based cohort study of 22 million individuals in the UK. Lancet. 2023;401:1878–1890. doi:10.1016/S0140-6736(23)00457-9

10. Jacobson DL, Gange SJ, Rose NR, Graham NM. Epidemiology and estimated population burden of selected autoimmune diseases in the United States. Clin Immunol Immunopathol. 1997;84:223–243. doi:10.1006/clin.1997.4412

11. Global burden of 288 causes of death and life expectancy decomposition in 204 countries and territories and 811 subnational locations, 1990-2021: a systematic analysis for the Global Burden of Disease Study 2021. Lancet. 2024;403:2100-2132.

12. Global age-sex-specific fertility, mortality, healthy life expectancy (HALE), and population estimates in 204 countries and territories, 1950-2019: a comprehensive demographic analysis for the Global Burden of Disease Study 2019. Lancet. 2020;396:1160-1203.

13. Murray CJL, Aravkin AY, Zheng P, et al. Global burden of 87 risk factors in 204 countries and territories, 1990–2019: a systematic analysis for the Global Burden of Disease Study 2019. Lancet. 2020;396:1223-1249. doi:10.1016/S0140-6736(20)30752-2

14. Global burden of 369 diseases and injuries in 204 countries and territories, 1990-2019: a systematic analysis for the Global Burden of Disease Study 2019. Lancet. 2020;396:1204-1222.

15. Boyle P, Parkin DM. Cancer registration: principles and methods. Statistical methods for registries. IARC Sci Publ. 1991:126–158.

16. Kim HJ, Fay MP, Feuer EJ, Midthune DN. Permutation tests for joinpoint regression with applications to cancer rates. Stat Med. 2000;19:335–351.

17. Global, regional, and national disability-adjusted life-years (DALYs) for 315 diseases and injuries and healthy life expectancy (HALE), 1990-2015: a systematic analysis for the Global Burden of Disease Study 2015. Lancet. 2016;388:1603-1658. doi:10.1016/S0140-6736(16)31460-X

18. Xie Y, Shi D, Wang X, Guan Y, Wu W, Wang Y. Prevalence trend and burden of neglected parasitic diseases in China from 1990 to 2019: findings from global burden of disease study. Front public Heal. 2023;11:1077723. doi:10.3389/fpubh.2023.1077723

19. Li R, Yuan X, Ou Y. Global burden of rheumatoid arthritis among adolescents and young adults aged 10-24 years: A trend analysis study from 1990 to 2019. PLoS One. 2024;19:e0302140. doi:10.1371/journal.pone.0302140

20. Gutierrez JP, Castañeda A, Agudelo-Botero M, Martínez-Valle A, Knight M, Lozano R. Performance evaluation of Mexico’s health system at the national and subnational level, 1990-2019: an analysis of the Health Access and Quality Index. Public Health. 2024;236:7-14. doi:10.1016/j.puhe.2024.07.009

21. Yu F, Chen H, Li Q, et al. Secular trend of mortality and incidence of rheumatoid arthritis in global ,1990–2019: an age period cohort analysis and joinpoint analysis. BMC Pulm Med. 2023;23:356. doi:10.1186/s12890-023-02594-2

22. Dadoun S, Zeboulon-Ktorza N, Combescure C, et al. Mortality in rheumatoid arthritis over the last fifty years: Systematic review and meta-analysis. Jt Bone Spine. 2013;80:29–33. 10.1016/j.jbspin.2012.02.005

23. Morales-Etchegaray I, Garcia-Carrasco M, Munguía-Realpozo P, et al. Changing trends in rheumatoid arthritis mortality in Mexico, from 1998 to 2017. Rheumatol Int. 2021;41:2225–2231. doi:10.1007/s00296-021-05013-z

24. Larrosa-Haro A, Abundis-Castro L, Contreras MB, et al. Epidemiologic trend of pediatric inflammatory bowel disease in Latin America: The Latin American Society for Pediatric Gastroenterology, Hepatology and Nutrition (LASPGHAN) Working Group. Rev Gastroenterol Mex. November 2020. doi:10.1016/j.rgmx.2020.07.010

25. Yamamoto-Furusho JK, Sarmiento-Aguilar A, Toledo-Mauriño JJ, et al. Incidence and prevalence of inflammatory bowel disease in Mexico from a nationwide cohort study in a period of 15 years (2000-2017). Medicine (Baltimore). 2019;98:e16291. doi:10.1097/MD.0000000000016291

26. Bouhuys M, Lexmond WS, van Rheenen PF. Pediatric Inflammatory Bowel Disease. Pediatrics. 2023;151. doi:10.1542/peds.2022-058037

27. Vuijk SA, Camman AE, de Ridder L. Considerations in Paediatric and Adolescent Inflammatory Bowel Disease. J Crohns Colitis. 2024;18:ii31-ii45. doi:10.1093/ecco-jcc/jjae087

28. Bertado-Cortés B, Venzor-Mendoza C, Rubio-Ordoñez D, et al. Demographic and clinical characterization of multiple sclerosis in Mexico: The REMEMBer study. Mult Scler Relat Disord. 2020;46:102575. doi:10.1016/j.msard.2020.102575

29. Lane J, Ng HS, Poyser C, Lucas RM, Tremlett H. Multiple sclerosis incidence: A systematic review of change over time by geographical region. Mult Scler Relat Disord. 2022;63:103932. doi:10.1016/j.msard.2022.103932

30. Wacher NH, Gómez-Díaz RA, Ascencio-Montiel I de J, Rascón-Pacheco RA, Aguilar-Salinas CA, Borja-Aburto VH. Type 1 diabetes incidence in children and adolescents in Mexico: Data from a nation-wide institutional register during 2000-2018. Diabetes Res Clin Pract. 2020;159:107949. doi:10.1016/j.diabres.2019.107949

31. Hernández-Garduño E. Asthma mortality among Mexican children: Rural and urban comparison and trends, 1999-2016. Pediatr Pulmonol. 2020;55:874-881. doi:10.1002/ppul.24658

32. Martínez-Martínez OA, Rodríguez-Brito A. Vulnerability in health and social capital: a qualitative analysis by levels of marginalization in Mexico. Int J Equity Health. 2020;19:24. doi:10.1186/s12939-020-1138-4

33. Phan K, Lee G, Fischer G. Pediatric psoriasis and association with cardiovascular and metabolic comorbidities: Systematic review and meta-analysis. Pediatr Dermatol. 2020;37:661–669. doi:10.1111/pde.14208

34. Brandi SL, Skov L, Strandberg-Larsen K, et al. Psoriasis and mental health in adolescents: A cross-sectional study within the Danish National Birth Cohort. J Affect Disord. 2024;358:318–325. doi:10.1016/j.jad.2024.05.009

