## Supplementary material for "Prevalence, Incidence, and Mortality of Autoimmune Diseases Among Adolescents and Young Adults in Mexico: An Analysis Based on the Global Burden of Disease Study 2021"

### Table S1. Projections of RA in adolescents and young adults in Mexico between 2022 and 2035

| **Year** | **ASPR (95% CI)** | **ASIR (95% CI)** | **ASMR (95% CI)** |
| --- | --- | --- | --- |
| 2022 | 46.08 (45.93–46.22) | 9.14 (9.1–9.17) | 0.02 (0.01–0.03) |
| 2023 | 46.48 (46.05–46.91) | 9.20 (9.11–9.29) | 0.02 (0.01–0.03) |
| 2024 | 46.87 (46.08–47.66) | 9.26 (9.1–9.43) | 0.02 (0.01–0.03) |
| 2025 | 47.25 (46.05–48.45) | 9.32 (9.07–9.57) | 0.02 (0.01–0.03) |
| 2026 | 47.63 (45.97–49.29) | 9.48 (9.04–9.83) | 0.02 (0.0–0.04) |
| 2027 | 48.0 (45.83–50.17) | 9.68 (9.07–10.29) | 0.02 (0.0–0.04) |
| 2028 | 48.36 (45.65–51.07) | 9.80 (9.00–10.59) | 0.02 (0.0–0.04) |
| 2029 | 48.72 (45.43–52.0) | 9.92 (8.92–10.91) | 0.02 (0.0–0.04) |
| 2030 | 49.07 (45.18–52.96) | 10.04 (8.92–11.25) | 0.02 (-0.0–0.04) |
| 2031 | 49.41 (44.89–53.93) | 10.16 (8.71–11.61) | 0.02 (-0.0–0.04) |
| 2032 | 50.21 (44.30–56.12) | 10.28 (8.58–11.98) | 0.02 (-0.0–0.04) |
| 2033 | 50.62 (43.89–57.35) | 10.40 (8.44–12.36) | 0.02 (-0.0–0.04) |
| 2034 | 51.03 (43.45–58.65) | 10.52 (8.29–12.76) | 0.02 (-0.01–0.05) |
| 2035 | 51.44 (42.97–59.92) | 10.64 (8.12–13.17) | 0.02 (-0.01–0.05) |

ASPR: Age-standardized prevalence rate; ASIR: Age-standardized incidence rate; ASMR: Age-standardized mortality rate. RA: rheumatoid arthritis.

Rates per 100,000 inhabitants.

### Table S2. Projections of IBD in adolescents and young adults in Mexico between 2022 and 2035

| **Year** | **ASPR (95% CI)** | **ASIR (95% CI)** | **ASMR (95% CI)** |
| --- | --- | --- | --- |
| 2022 | 0.76 (0.74–0.78) | 0.11 (0.10–0.12) | 0.04 (0.03–0.05) |
| 2023 | 0.76 (0.72–0.80) | 0.11 (0.10–0.12) | 0.04 (0.03–0.05) |
| 2024 | 0.76 (0.70–0.82) | 0.11 (0.10–0.12) | 0.04 (0.03–0.05) |
| 2025 | 0.76 (0.69–0.83) | 0.11 (0.09–0.13) | 0.04 (0.03–0.05) |
| 2026 | 0.76 (0.67–0.85) | 0.11 (0.09–0.13) | 0.04 (0.03–0.05) |
| 2027 | 0.76 (0.65–0.87) | 0.11 (0.09–0.13) | 0.04 (0.03–0.05) |
| 2028 | 0.76 (0.64–0.88) | 0.11 (0.09–0.13) | 0.04 (0.02–0.06) |
| 2029 | 0.76 (0.63–0.89) | 0.11 (0.09–0.13) | 0.04 (0.02–0.06) |
| 2030 | 0.76 (0.61–0.91) | 0.11 (0.08–0.14) | 0.04 (0.02–0.06) |
| 2031 | 0.76 (0.60–0.92) | 0.11 (0.08–0.14) | 0.04 (0.02–0.06) |
| 2032 | 0.76 (0.59–0.93) | 0.11 (0.08–0.14) | 0.04 (0.02–0.06) |
| 2033 | 0.76 (0.58–0.94) | 0.11 (0.08–0.14) | 0.04 (0.02–0.06) |
| 2034 | 0.76 (0.57–0.95) | 0.11 (0.08–0.14) | 0.04 (0.02–0.06) |
| 2035 | 0.76 (0.56–0.96) | 0.11 (0.08–0.14) | 0.04 (0.02–0.06) |

ASPR: Age-standardized prevalence rate; ASIR: Age-standardized incidence rate; ASMR: Age-standardized mortality rate. IBD: inflammatory bowel disease.

Rates per 100,000 inhabitants.

### Table S3. Projections of MS in adolescents and young adults in Mexico between 2022 and 2035

| **Year** | **ASPR (95% CI)** | **ASIR (95% CI)** | **ASMR (95% CI)** |
| --- | --- | --- | --- |
| 2022 | 2.02 (2.00–2.03) | 0.39 (0.38–0.40) | 0.03 (0.03–0.04) |
| 2023 | 2.02 (1.99–2.05) | 0.39 (0.37–0.41) | 0.03 (0.02–0.04) |
| 2024 | 2.03 (1.98–2.07) | 0.39 (0.37–0.42) | 0.03 (0.02–0.04) |
| 2025 | 2.03 (1.97–2.10) | 0.39 (0.36–0.43) | 0.03 (0.02–0.04) |
| 2026 | 2.04 (1.95–2.12) | 0.40 (0.36–0.43) | 0.03 (0.02–0.04) |
| 2027 | 2.04 (1.93–2.15) | 0.40 (0.35–0.44) | 0.03 (0.02–0.05) |
| 2028 | 2.04 (1.92–2.17) | 0.40 (0.35–0.44) | 0.03 (0.02–0.05) |
| 2029 | 2.05 (1.90–2.20) | 0.40 (0.34–0.45) | 0.04 (0.02–0.05) |
| 2030 | 2.05 (1.88–2.23) | 0.40 (0.34–0.45) | 0.04 (0.02–0.05) |
| 2031 | 2.06 (1.86–2.26) | 0.40 (0.34–0.46) | 0.04 (0.02–0.05) |
| 2032 | 2.06 (1.83–2.29) | 0.41 (0.35–0.46) | 0.04 (0.02–0.05) |
| 2033 | 2.07 (1.81–2.32) | 0.41 (0.35–0.47) | 0.04 (0.02–0.05) |
| 2034 | 2.07 (1.79–2.35) | 0.41 (0.34–0.47) | 0.04 (0.02–0.06) |
| 2035 | 2.07 (1.76–2.39) | 0.41 (0.34–0.48) | 0.04 (0.02–0.06) |

ASPR: Age-standardized prevalence rate; ASIR: Age-standardized incidence rate; ASMR: Age-standardized mortality rate. MS: multiple sclerosis.

Rates per 100,000 inhabitants.

**Table S4. Projections of T1DM in adolescents and young adults in Mexico between 2022 and 2035**

| **Year** | **ASPR (95% CI)** | **ASIR (95% CI)** | **ASMR (95% CI)** |
| --- | --- | --- | --- |
| 2022 | 179.04 (171.62–186.46) | 10.03 (9.86–10.21) | 0.52 (0.49–0.56) |
| 2023 | 170.95 (154.35–187.55) | 9.82 (9.46–10.18) | 0.53 (0.47–0.58) |
| 2024 | 162.86 (135.09–190.63) | 9.61 (9.03–10.19) | 0.53 (0.47–0.60) |
| 2025 | 154.77 (114.11–195.43) | 9.40 (8.57–10.22) | 0.54 (0.46–0.61) |
| 2026 | 146.68 (91.63–201.73) | 9.19 (8.09–10.28) | 0.54 (0.46–0.63) |
| 2027 | 138.59 (67.78–209.40) | 8.97 (7.58–10.37) | 0.55 (0.45–0.64) |
| 2028 | 130.50 (42.67–218.33) | 8.76 (7.05–10.48) | 0.55 (0.45–0.65) |
| 2029 | 122.41 (16.39–228.43) | 8.55 (6.49–10.61) | 0.56 (0.45–0.66) |
| 2030 | 114.32 (-11.00–239.64) | 8.34 (5.92–10.76) | 0.56 (0.45–0.68) |
| 2031 | 106.23 (-39.42–251.88) | 8.13 (5.32–10.93) | 0.57 (0.44–0.69) |
| 2032 | 98.14 (-68.84–265.12) | 7.91 (4.71–11.12) | 0.57 (0.44–0.70) |
| 2033 | 90.05 (-99.20–279.30) | 7.70 (4.08–11.32) | 0.57 (0.44–0.71) |
| 2034 | 81.96 (-130.48–294.40) | 7.49 (3.44–11.54) | 0.58 (0.44–0.72) |
| 2035 | 73.87 (-162.62–310.36) | 7.28 (2.78–11.78) | 0.58 (0.44–0.73) |

ASPR: Age-standardized prevalence rate; ASIR: Age-standardized incidence rate; ASMR: Age-standardized mortality rate. T1DM: Type 1 diabetes mellitus.

Rates per 100,000 inhabitants.

### Table S5. Projections of Asthma in adolescents and young adults in Mexico between 2022 and 2035

| **Year** | **ASPR (95% CI)** | **ASIR (95% CI)** | **ASMR (95% CI)** |
| --- | --- | --- | --- |
| 2022 | 3092.27 (3034.98–3149.56) | 460.85 (456.49–465.21) | 0.18 (0.15–0.20) |
| 2023 | 3111.88 (2983.78–3239.98) | 461.17 (451.42–470.92) | 0.17 (0.14–0.21) |
| 2024 | 3131.49 (2917.14–3345.84) | 461.49 (445.17–477.81) | 0.17 (0.12–0.21) |
| 2025 | 3151.10 (2837.32–3464.87) | 461.81 (437.92–485.70) | 0.16 (0.11–0.21) |
| 2026 | 3170.71 (2745.85–3595.56) | 462.13 (429.78–494.48) | 0.16 (0.10–0.21) |
| 2027 | 3190.31 (2643.83–3736.80) | 462.45 (420.84–504.06) | 0.15 (0.09–0.21) |
| 2028 | 3209.92 (2532.10–3887.75) | 462.77 (411.16–514.38) | 0.15 (0.08–0.21) |
| 2029 | 3229.53 (2411.31–4047.75) | 463.09 (400.79–525.39) | 0.14 (0.07–0.21) |
| 2030 | 3249.14 (2282.03–4216.25) | 463.41 (389.77–537.05) | 0.14 (0.07–0.21) |
| 2031 | 3268.75 (2144.70–4392.80) | 463.73 (378.14–549.32) | 0.13 (0.06–0.21) |
| 2032 | 3288.36 (1999.73–4576.99) | 464.05 (365.93–562.17) | 0.13 (0.05–0.21) |
| 2033 | 3307.97 (1847.44–4768.50) | 464.37 (353.16–575.58) | 0.13 (0.04–0.21) |
| 2034 | 3327.58 (1688.14–4967.02) | 464.69 (339.86–589.52) | 0.12 (0.03–0.21) |
| 2035 | 3347.19 (1522.09–5172.29) | 465.01 (326.05–603.98) | 0.12 (0.02–0.21) |

ASPR: Age-standardized prevalence rate; ASIR: Age-standardized incidence rate; ASMR: Age-standardized mortality rate.

Rates per 100,000 inhabitants.

**Table S6. Projections of Psoriasis in adolescents and young adults in Mexico between 2022 and 2035**

| **Year** | **ASPR (95% CI)** | **ASIR (95% CI)** |
| --- | --- | --- |
| 2022 | 467.71 (467.12–468.30) | 57.23 (57.10–57.35) |
| 2023 | 467.69 (466.38–469.00) | 57.15 (56.79–57.51) |
| 2024 | 467.67 (465.47–469.87) | 57.08 (56.42–57.73) |
| 2025 | 467.65 (464.43–470.87) | 57.00 (56.00–58.00) |
| 2026 | 467.63 (463.28–471.98) | 56.93 (55.54–58.32) |
| 2027 | 467.61 (462.01–473.21) | 56.86 (55.04–58.68) |
| 2028 | 467.59 (460.64–474.54) | 56.78 (54.49–59.07) |
| 2029 | 467.57 (459.19–475.96) | 56.71 (53.92–59.50) |
| 2030 | 467.55 (457.64–477.46) | 56.64 (53.31–59.96) |
| 2031 | 467.53 (456.01–479.05) | 56.56 (52.67–60.45) |
| 2032 | 467.51 (454.30–480.72) | 56.49 (52.01–60.97) |
| 2033 | 467.49 (452.52–482.46) | 56.41 (51.31–61.52) |
| 2034 | 467.47 (450.67–484.27) | 56.34 (50.59–62.09) |
| 2035 | 467.45 (448.75–486.15) | 56.27 (49.85–62.68) |
